# The impact of pre-stroke statin use on baseline corrected infarct volume and collateral perfusion

**DOI:** 10.64898/2026.06.09.26355321

**Authors:** Kirsten G. Coupland, Barbara Toson, Kristy Martin, Tom Lillicrap, Christopher R. Levi, Alex Pinheiro, Carlos Garcia-Esperon, Neil J. Spratt

## Abstract

1.0

Stroke is a leading cause of disability and mortality worldwide, with ischaemic stroke the most prevalent type. Statins, used for cholesterol management, have demonstrated benefits in reducing stroke risk and improving outcomes in preclinical studies. However, the impact of pre-stroke statin use on stroke outcomes remain inconsistent. In this study, we aim to evaluate whether pre-stroke statin use is associated with greater volume of salvaged tissue and improved cerebral collateral perfusion.

A retrospective analysis was conducted using data from 281 patients presenting with acute ischemic stroke to the John Hunter Hospital between May 2015 and May 2020. Patients were grouped based on pre-stroke statin use, and clinical variables, including infarct volume and collateral perfusion, were assessed. The primary outcome was salvage volume derived from baseline perfusion lesion volume minus infarct volume at follow-up. Collateral perfusion was measured by the hypoperfusion volume defined by delay time (DT)>6 seconds divided by the hypoperfusion volume defined by DT >2 seconds.

Patients on statins at admission were significantly older and had more comorbidities. No significant association was found between pre-stroke statin use and salvage volume or collateral perfusion after adjusting for covariates. Larger initial infarct core was a significant predictor of salvage volume due to larger salvageable tissue volume at baseline. These findings indicate that pre-morbid statin use is not associated with larger salvage volume or improved cerebral collateral perfusion.

**Highlights:**

- There is pre-clinical and clinical evidence that pre-stroke statin use is protective but randomised controlled trials (RCTs) have not replicated these findings.
- Performed retrospective analysis of salvage volume in stroke patients with and without pre-morbid statin use.
- Pre-stroke statin use was not associated with greater salvage volume or improved cerebral collateral perfusion.
- Baseline infarct volume was the most significant predictor of salvage volume due to those with a larger initial infarct core having a greater volume of hypoperfused tissue.

## 4.0 Introduction

Stroke is the third-leading cause of disability and death worldwide with 143 million disability adjusted life years lost due to the disease [1]. Ischaemic stroke is the most common form, accounting for ∼80% of all strokes. Statins reduce the likelihood of primary and secondary stroke events, and there is also evidence that they reduce stroke volume in experimental models and improve outcomes in clinical studies. However findings are inconsistent and randomised clinical trials (RCTs) have not found a significant benefit [2–4].

Statins inhibit HMG-CoA (3-hydroxy-3-methyl-glutaryl-coenzyme A) reductase, the rate-limiting enzyme for cholesterol synthesis, and are used as an effective treatment for hypercholesterolaemia. In addition to their capacity to reduce cholesterol levels, a systematic review and meta-analysis identified a 25.12% reduction in infarct volume and improved neurological score in experimental stroke models. The effect was particularly prominent in animals given statins prior to stoke [5]. This appears to fit with findings from patient cohorts in which patients on statins prior to stroke have better functional outcomes and lower likelihood of all-cause death at 30 and 90 days compared to those not on statins [4,6]. However, RCTs in which patients were randomised to statin use after the stroke event did not demonstrate any beneficial effect on stroke outcome [4]. Together, this indicates that being on statins prior to a stroke may confer protective benefits and this may be part of the reason why RCTs of statin dosing in the acute phase of stroke have not found a significant improvement.

An important predictor of stroke outcome are the leptomeningeal collateral vessels. Leptomeningeal collateral vessels link distal arteriolar branches and supply watershed regions between the cerebral arteries [7,8]. In the event of an ischaemic stroke, collateral vessels provide an alternate route for blood flow towards the site of decreased perfusion. This sustains the salvageable penumbra, with a higher collateral grade being associated with smaller infarct volume and better stroke outcomes [9]. Several studies indicate that statins promote proliferation, maturation, and migration of endothelial cells in a manner indicative of angiogenesis [10–13]. In addition, pre-stroke statin use has been reported to correlate with better collaterals [14–17]. As such, improved leptomeningeal collateral grade, and therefore perfusion, may be part of the mechanism by which premorbid statin use improves stroke outcome.

We hypothesise that statin use prior to ischaemic stroke stimulates collateral angiogenesis, thus improving collateral grade and blood flow, resulting in greater salvage volume for the same initially ischaemic territory. If prophylactic prescription of statins prior to stroke is associated with smaller final infarct volume this may inform future prescription practices in populations at high risk of stroke. Using historical patient data collected by the Stroke Unit at the John Hunter Hospital (Newcastle, Australia) between May 2015 and May 2020, we assessed whether statins are associated with better collateral perfusion relative to patients not on statins, and whether pre-stroke statin use is associated with larger salvage volume when normalised to perfusion lesion volume at admission.

## 5.0 Methods

### 5.1 Participants

We retrospectively analysed data from acute ischaemic stroke patients enrolled in the Targeting Optimal Thrombolysis (TOTO) cohort presenting to the John Hunter Hospital (New South Wales, Australia) from May 2015 to May 2020 [18]. Patients eligible for inclusion were aged ≥18 years with clinicoradiological evidence of acute brain ischemia who have multimodal imaging using either computed tomography (CT) or magnetic resonance imaging (MRI) at the baseline evaluation (i.e., including angiography and perfusion imaging), with no evidence of intracranial haemorrhage, stroke mimic or other non-stroke pathology. Patients were required to have follow-up CT or magnetic resonance perfusion imaging performed between 18-36 hours after hospital presentation.

Patients provided written informed consent for their clinical, imaging and blood sample data to be used for research. All patients were provided with standard acute stroke unit care. The study received approval from the Hunter New England Local Health District Human Research Ethics Committee (reference 2019/ETH03892). All acquired data was de-identified, stored electronically and password protected.

### 5.2 Assessment

Patients were divided into two groups: statin use prior to hospital admission, or no statin use prior to hospital admission. Clinical variables that are recognised predictors of stroke outcome were collected, including pre-stroke modified Rankin Score (mRS), age, sex, stroke severity (measured using the National Institutes of Health Stroke Scale [NIHSS]), blood glucose, blood pressure, concomitant antithrombotic therapy and smoking status. NIHSS follows the scoring system: 0: no signs of stroke, 1-5: mild stroke, 5-14: mild to moderately severe, 15-24: severe, >25: very severe. Treatment-related variables included use of thrombolytics and/or endovascular thrombectomy (EVT) and success of recanalization, scored using the thrombolysis in cerebral infarction (TICI) score. Disease and medication variables including diabetes, hypertension, atrial fibrillation, and hypercholesterolaemia were collected.

We collected core and penumbra volumes at baseline and follow-up imaging for each group, and calculated salvage volume (volume of hypoperfused tissue [delay time>4] minus infarct core volume at follow-up [diffusion weighted imaging, DWI]). Using contrast peak time delay and contrast peak density it is possible to derive a quantified estimate of CT perfusion collateral index to assess differences in collateral perfusion as previously reported [9]. This was correlated to the patient’s pre-stroke medication status.

### 5.3 Neuroimaging

All patients underwent baseline: non-contrast CT, CT perfusion (CTP) and CT angiogram (CTA) or multimodal MRI (diffusion, T2, T1, perfusion and MR angiography), and post-perfusion therapy MRI or CT. The latter was performed 18-36 hours after admission. Patients who underwent EVT had preprocedural and postprocedural angiograms. Imaging was performed as previously outlined [18]. Briefly, CT imaging was derived from a 320-slice Aquilion ONE scanner (Canon medical systems, Otawara, Japan). Image scanning started 7s after intravenous injection (40 mL at 6 mL/s) of non-ionic iodinated contrast (Ultravist 370; Bayer HealthCare, Berlin, Germany). Imaging lasted for 60s, with acquisition at 19 images per slice. Magnetic resonance perfusion imaging was performed on a 1.5-Tesla scanner (Siemens Avanto, Erlangen, Germany) and included axial isotropic diffusion weighted imaging (DWI), echoplanar spin-echo sequence, time of flight magnetic resonance angiography, and bolus-tracking perfusion weighted imaging. Following a bolus of gadolinium contrast (Magnevist; Bayer HealthCare) into the antecubital vein (0.2 mmol/kg at 5 mL/s), perfusion images were obtained within acquisition parameters. The scanning lasted for 60s, resulting in 40 images per slice. A total of 19 slices were obtained.

CTP and MR perfusion imaging were post-processed using the commercial software MIStar (Apollo Medical Imaging Technology, Melbourne, Australia) to measure volumes of the ischaemic core, penumbra, and severe hypoperfusion, and CT perfusion collateral index. Penumbra was defined as the tissue with a delay time (DT) ≥4s and relative cerebral blood flow (CBF) ≥30% of the contralateral hemisphere using either CTP or magnetic resonance perfusion in MIStar. Ischaemic core was defined as the tissue with a DT≥3s and a relative CBF≤30% of the contralateral hemisphere on CTP or an apparent diffusion coefficient threshold of <620 x 10^-6^mm^2^/s on MRI-DWI. All images were reprocessed and analysed using the same version of the MIStar software.

### 5.4 CT perfusion collateral index

CT perfusion collateral index was determined as outlined previously [9]. Briefly, collateral status was assessed on CTP, measuring the ratio of severely hypoperfused tissue within any region of hypoperfusion. The following equation was used to define CT perfusion collateral index: CT perfusion collateral index = hypoperfusion volume of DT>6 seconds/hypoperfusion volume of DT>2 seconds. DT>6 seconds measures the severely hypoperfused region without collateral flow, whereas DT>2 seconds defines the region with any hypoperfusion on the CTP. The CT perfusion collateral index is a continuous variable, with a higher value representing a larger region without collateral flow and is indicative of poorer collateral flow.

### 5.5 Statistical analysis

Baseline characteristics by statin use (none or pre-stroke) were summarized by counts or percentages for categorical variables, means with standard deviations (SDs) for all continuous variables and additional median and interquartile range for non-normally distributed continuous variables. We compared means and prevalences of baseline characteristics using absolute standardised mean differences (SMD), with a threshold of 0.10 indicating imbalance. Salvage volume was calculated as baseline DT>4 minus follow-up DWI volume. Penumbra volume was calculated as DT>4 minus CBF<30%. Smoking status was categorised as never, former, or current smoker; respondents who did not provide an answer were assigned to a separate ‘Not answered’ category to retain these participants in the analysis without introducing additional missingness in complete-case analysis.

Unadjusted comparisons for CBF<30%, salvage volume, and CT perfusion collateral index were performed using Wilcoxon rank-sum test. To mitigate the influence of outliers and violations of normality and homoscedasticity assumptions, salvage volume was analysed using a robust regression with iteratively reweighted least squares method. A subpopulation analysis was performed on patients who underwent EVT and for whom a TICI score was available (n=86) to examine the effect of successful vessel recanalization on the relationship between statin use on admission and salvage volume. TICI scores were categorised into poor-moderate (0-2b) and excellent (2c-3) recanalization. Cerebral perfusion at DT>2 and DT>6 was reported using geometric means and 95% CIs. The CT perfusion collateral index was analysed using a fractional logistic regression to assess the effects of statin on the index. All baseline characteristics with SMD above the threshold were included in the models to adjust for potential confounders.

The primary analysis was performed on a complete-case population. To address potential bias arising from missing data, sensitivity analyses using multiple imputation by chained equations (MICE) was performed. Imputations were performed separately within the whole study population or within the relevant analytic subsets to ensure that the imputed values reflected the distributions and associations specific to each analysis sample. Binary variables were imputed using logistic regression, and categorical variables using multinomial logistic regression. All variables included in the analysis model were included in the imputation model, and no auxiliary variables were used. Estimates from 20 imputed datasets were pooled using Rubin’s rules. All analyses were performed using Stata version 18 (StataCorp, College Station, TX).

## 6.0 Results

### 6.1 Patient Demographics

From an initial database of 322 patients, we excluded patients with missing critical data (follow-up imaging n=38, incorrect patient ID n=1, second stroke n=1, missing age and stroke onset date n=1). Our final data set examined 281 patients. There were 105 patients on statins at time of admission and 176 patients not on statins at admission. Baseline demographics and clinical characteristics were compared by statin use (Table 1). There were 181 men and 100 women, with a median age of 71 years (interquartile range, 58 to 78). The mean baseline NIHSS score was 10.78 (SD ±6.47). Of the patients on statins prior to stroke for whom details of the prescribed statin were available (n=69), 92% were prescribed atorvastatin. Numbers were insufficient for grouped analysis by specific statin and thus patients on statins prior to stroke were pooled for subsequent analysis.

**Table 1.**
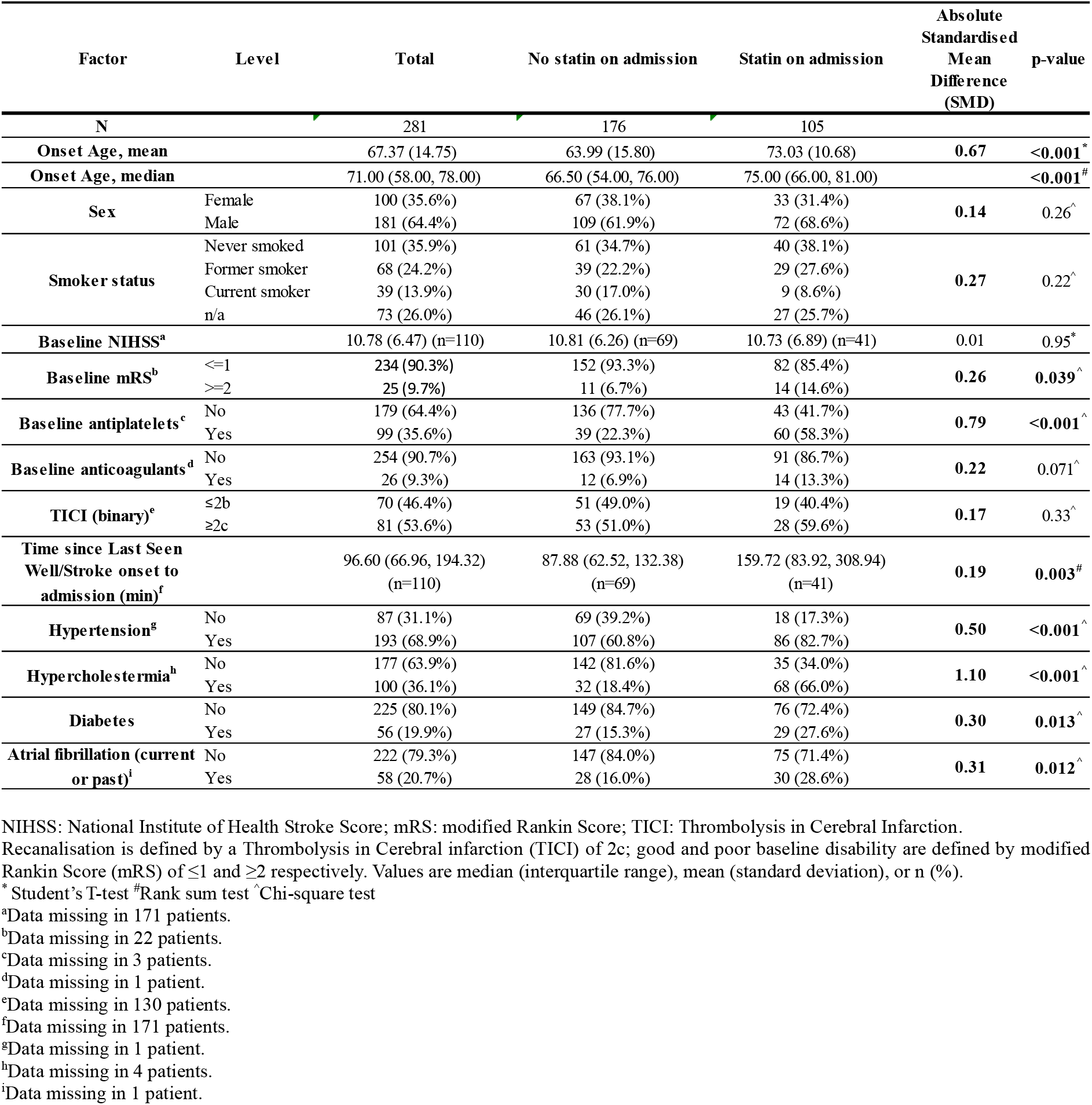
Patient characteristics by statin use on admission and absolute standardized mean differences (SMD).

| Factor | Level | Total | No statin on admission | Statin on admission | Absolute Standardised Mean Difference (SMD) | p-value |
| --- | --- | --- | --- | --- | --- | --- |
| N |  | 281 | 176 | 105 |  |  |
| Onset Age, mean |  | 67.37 (14.75) | 63.99 (15.80) | 73.03 (10.68) | 0.67 | <0.001* |
| Onset Age, median |  | 71.00 (58.00, 78.00) | 66.50 (54.00, 76.00) | 75.00 (66.00, 81.00) |  | <0.001# |
| Sex | Female | 100 (35.6%) | 67 (38.1%) | 33 (31.4%) | 0.14 | 0.26^ |
|  | Male | 181 (64.4%) | 109 (61.9%) | 72 (68.6%) |  |  |
| Smoker status | Never smoked | 101 (35.9%) | 61 (34.7%) | 40 (38.1%) | 0.27 | 0.22^ |
|  | Former smoker | 68 (24.2%) | 39 (22.2%) | 29 (27.6%) |  |  |
|  | Current smoker | 39 (13.9%) | 30 (17.0%) | 9 (8.6%) |  |  |
|  | n/a | 73 (26.0%) | 46 (26.1%) | 27 (25.7%) |  |  |
| Baseline NIHSS<sup>a</sup> |  | 10.78 (6.47) (n=110) | 10.81 (6.26) (n=69) | 10.73 (6.89) (n=41) | 0.01 | 0.95* |
| Baseline mRS<sup>b</sup> | ≤1 | 234 (90.3%) | 152 (93.3%) | 82 (85.4%) | 0.26 | 0.039^ |
|  | ≥2 | 25 (9.7%) | 11 (6.7%) | 14 (14.6%) |  |  |
| Baseline antiplatelets<sup>c</sup> | No | 179 (64.4%) | 136 (77.7%) | 43 (41.7%) | 0.79 | <0.001^ |
|  | Yes | 99 (35.6%) | 39 (22.3%) | 60 (58.3%) |  |  |
| Baseline anticoagulants<sup>d</sup> | No | 254 (90.7%) | 163 (93.1%) | 91 (86.7%) | 0.22 | 0.071^ |
|  | Yes | 26 (9.3%) | 12 (6.9%) | 14 (13.3%) |  |  |
| TICI (binary)<sup>e</sup> | ≤2b | 70 (46.4%) | 51 (49.0%) | 19 (40.4%) | 0.17 | 0.33^ |
|  | ≥2c | 81 (53.6%) | 53 (51.0%) | 28 (59.6%) |  |  |
| Time since Last Seen Well/Stroke onset to admission (min)<sup>f</sup> |  | 96.60 (66.96, 194.32) (n=110) | 87.88 (62.52, 132.38) (n=69) | 159.72 (83.92, 308.94) (n=41) | 0.19 | 0.003# |
| Hypertension<sup>g</sup> | No | 87 (31.1%) | 69 (39.2%) | 18 (17.3%) | 0.50 | <0.001^ |
|  | Yes | 193 (68.9%) | 107 (60.8%) | 86 (82.7%) |  |  |
| Hypercholestermia<sup>h</sup> | No | 177 (63.9%) | 142 (81.6%) | 35 (34.0%) | 1.10 | <0.001^ |
|  | Yes | 100 (36.1%) | 32 (18.4%) | 68 (66.0%) |  |  |
| Diabetes | No | 225 (80.1%) | 149 (84.7%) | 76 (72.4%) | 0.30 | 0.013^ |
|  | Yes | 56 (19.9%) | 27 (15.3%) | 29 (27.6%) |  |  |
| Atrial fibrillation (current or past)<sup>i</sup> | No | 222 (79.3%) | 147 (84.0%) | 75 (71.4%) | 0.31 | 0.012^ |
|  | Yes | 58 (20.7%) | 28 (16.0%) | 30 (28.6%) |  |  |

NIHSS: National Institute of Health Stroke Score; mRS: modified Rankin Score; TICI: Thrombolysis in Cerebral Infarction.
Recanalisation is defined by a Thrombolysis in Cerebral infarction (TICI) of 2c; good and poor baseline disability are defined by modified Rankin Score (mRS) of ≤1 and ≥2 respectively. Values are median (interquartile range), mean (standard deviation), or n (%).
\* Student's T-test #Rank sum test ^Chi-square test
<sup>a</sup>Data missing in 171 patients.
<sup>b</sup>Data missing in 22 patients.
<sup>c</sup>Data missing in 3 patients.
<sup>d</sup>Data missing in 1 patient.
<sup>e</sup>Data missing in 130 patients.
<sup>f</sup>Data missing in 171 patients.
<sup>g</sup>Data missing in 1 patient.

Patients on statins were older, had poorer functional status, were on more concurrent medications, presented to hospital later from symptom onset, and had higher levels of all measured stroke risk factors than those not on statins at admission (Table 1). All baseline characteristics were imbalanced between the two groups, with the exception of baseline NIHSS score.

### 6.2 Being on statins at admission is not significantly associated with larger salvage volume

Unadjusted analyses showed no statistically significant differences in salvage volume, acute core volume (CBF<30%), CT perfusion collateral index, or follow-up infarct volume (DWI) between patients who were and were not on statins at admission. The point estimate of median salvage volume was not significantly different between statin groups (p = 0.839, Table 2).

**Table 2.** Unadjusted comparisons of imaging-based outcome measures by statin use at admission.

|  | Statin on admission |  | p-value |
| --- | --- | --- | --- |
|  | No | Yes |  |
| N | 176 | 105 |  |
| Salvage volume (mL) | 1.23 (-2.79 – 37.51) | 1.9 (-3.02 – 35.90) | 0.839 |
| Acute core volume (mL) | 5.00 (0.00 – 19.50) | 3.00 (0.00 – 15.00) | 0.180 |
| CT perfusion collateral index | 0.14 (0.16) | 0.14 (0.16) | 0.700 |
| Follow-up infarct volume (median, mL) | 8.55 (2.29 – 33.12) | 9.60 (2.10 – 24.10) | 0.720 |
| Follow-up infarct volume (mean, mL) | 27.36 (45.15) | 20.23 (27.14) | 0.140 |

Acute core volume is measured by cerebral blood flow (CBF) <30% on CTP; CT perfusion collateral index is calculated by the hypoperfusion volume defined by delay time (DT)>6 seconds divided by the hypoperfusion volume defined by DT>2 seconds; follow-up infarct volume is measured by diffusion-weighted imaging at 18-36 hours after admission. Values are median (interquartile range) or mean (standard deviation).

Robust regression analysis similarly indicated that statin use at admission had no effect on salvage volume (b = –4.32, 95% CI: –14.26−5.62, p = 0.393; Table 3). Acute core volume (CBF<30%) was the dominant predictor of salvage volume (b = 0.81, 95% CI: 0.64 to 0.99, p <0.001). Subsequent analyses identified that this is due to penumbra volume correlating with CBF<30% (i.e., the larger the stroke core, the larger the penumbra and thus the more tissue you have to salvage; Supplementary Table S1). Although Baseline mRS does not meet the conventional threshold for significance (b = 14.12, 95% CI: –0.41 - 28.66, p = 0.057), patients with worse pre-stroke disability (mRS≥2) tended to have a greater salvage volume. Smoker status, diabetes, hypertension, hypercholesterolemia, baseline anticoagulant and/or antiplatelet use, and atrial fibrillation did not show a significant association with salvage volume in this model (Table 3). Results from sensitivity analysis following multiple imputation consistently confirmed that statin use at admission was not associated with salvage volume, while acute core volume was the dominant predictor (Supplementary Table S2). In addition, being a former smoker was negatively associated with salvage volume (b = -10.31, 95% CI -20.23 - -0.39, p = 0.042).

**Table 3.** Robust regression coefficients for predictors of salvage volume (N=250^*^).

|  | **b Coefficient** | **95% confidence interval** | **p-value** |
| --- | --- | --- | --- |
| **Statin on admission** | -4.32 | -14.26 - 5.62 | 0.393 |
| **Acute core volume (mL)** | 0.81 | 0.64 - 0.99 | **<0.001** |
| **Age at onset** | -0.14 | -0.49 - 0.21 | 0.436 |
| **Sex** |  |  |  |
| Male | 3.21 | -5.31 - 11.74 | 0.458 |
| **Smoker status (reference Never smoked)** |  |  | 0.278 |
| Former smoker | -9.35 | -19.96 - 1.26 | 0.084 |
| Current smoker | -9.06 | -22.39 - 4.27 | 0.182 |
| Not answered | -2.86 | -13.16 - 7.43 | 0.584 |
| **Baseline mRS $\geq 2$** | 14.12 | -0.41 - 28.66 | 0.057 |
| **Baseline anticoagulants** | 8.03 | -8.65 - 24.71 | 0.344 |
| **Baseline antiplatelets** | 1.81 | -7.90 - 11.51 | 0.714 |
| **Diabetes** | -0.12 | -10.96 - 10.72 | 0.983 |
| **Hypercholestermia** | 3.31 | -6.65 - 13.27 | 0.514 |
| **Hypertension** | 4.68 | -6.00 - 15.37 | 0.389 |
| **Atrial Fibrillation** | -0.95 | -12.80 - 10.91 | 0.875 |

mRS: modified Rankin Score; Acute core volume is measured by cerebral blood flow <30% on CTP.
\*Model with imputed values for missing data in Supplementary Table 2.

In patients who underwent EVT, statin use at admission was also not significantly associated with salvage volume after adjusting for covariates (b = 8.91, 95% CI –27.28 to 45.09; p = 0.625; Table 4). Similarly, the TICI coefficient was not significantly associated with salvage volume (b = 14.56, 95% CI –16.30 to 45.42; p = 0.350). In contrast, acute core volume (CBF<30%) was significantly associated with salvage volume (b = 0.77, 95% CI 0.20 to 1.34; p = 0.009). No other covariates were significantly associated with salvage volume in the EVT-treated cohort. Sensitivity analysis confirmed that statin use at admission and TICI were not significantly associated with salvage volume (Supplementary Table 3). The magnitude effect of acute core volume (CBF<30%) was attenuated but remained significant (b = 0.46, 95% CI 0.02 to 0.90; p = 0.029), while antiplatelet use was associated with a significant increase in salvage volume (b = 40.15, 95% CI 6.17 to 74.12; p = 0.020).

**Table 4.**
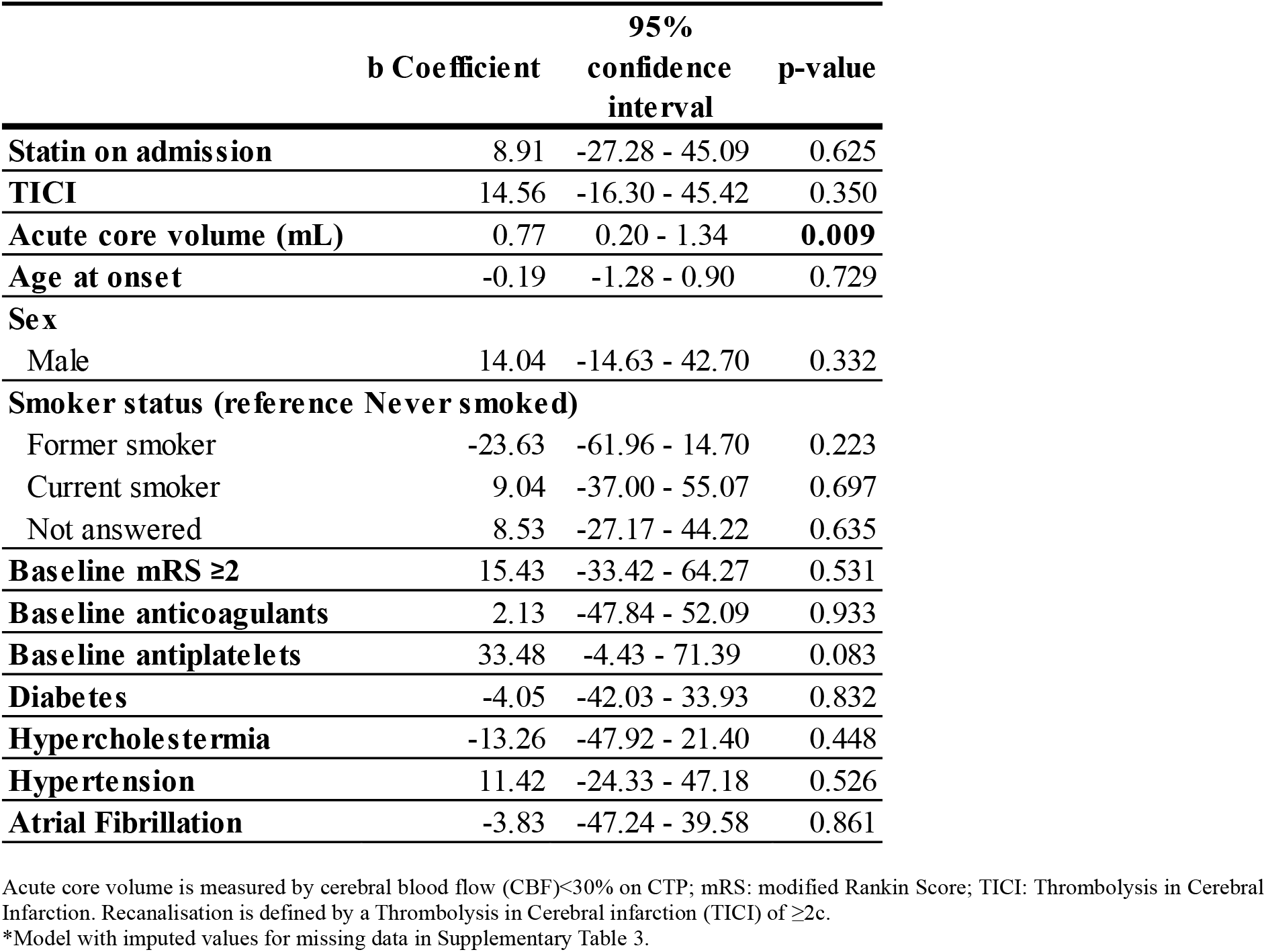
Robust regression coefficients for predictors of salvage volume in EVT subpopulation (N=86^*^)

|  | **b Coefficient** | **95% confidence interval** | **p-value** |
| --- | --- | --- | --- |
| **Statin on admission** | 8.91 | -27.28 - 45.09 | 0.625 |
| **TICI** | 14.56 | -16.30 - 45.42 | 0.350 |
| **Acute core volume (mL)** | 0.77 | 0.20 - 1.34 | **0.009** |
| **Age at onset** | -0.19 | -1.28 - 0.90 | 0.729 |
| **Sex** |  |  |  |
| Male | 14.04 | -14.63 - 42.70 | 0.332 |
| **Smoker status (reference Never smoked)** |  |  |  |
| Former smoker | -23.63 | -61.96 - 14.70 | 0.223 |
| Current smoker | 9.04 | -37.00 - 55.07 | 0.697 |
| Not answered | 8.53 | -27.17 - 44.22 | 0.635 |
| **Baseline mRS $\geq 2$** | 15.43 | -33.42 - 64.27 | 0.531 |
| **Baseline anticoagulants** | 2.13 | -47.84 - 52.09 | 0.933 |
| **Baseline antiplatelets** | 33.48 | -4.43 - 71.39 | 0.083 |
| **Diabetes** | -4.05 | -42.03 - 33.93 | 0.832 |
| **Hypercholestermia** | -13.26 | -47.92 - 21.40 | 0.448 |
| **Hypertension** | 11.42 | -24.33 - 47.18 | 0.526 |
| **Atrial Fibrillation** | -3.83 | -47.24 - 39.58 | 0.861 |

Acute core volume is measured by cerebral blood flow (CBF)<30% on CTP; mRS: modified Rankin Score; TICI: Thrombolysis in Cerebral Infarction. Recanalisation is defined by a Thrombolysis in Cerebral infarction (TICI) of $\geq 2$ .
\*Model with imputed values for missing data in Supplementary Table 3.

### 6.3 Being on statins at admission is not associated with improved collateral perfusion

We next examined whether the CT perfusion collateral index, calculated by the hypoperfusion volume defined by DT>6 seconds divided by the hypoperfusion volume defined by DT>2 seconds, in patients for whom the volume of tissue with CBF<30% was greater than 15 mL (n=72) differed between patients on statins at admission and those not on statins at admission [9,20]. Statin use on admission was not a significant predictor of the CT perfusion collateral index (OR 0.87, 95% CI 0.59-1.29, p=0.493; Table 5). Sex was the only variable with a statistically significant association to the CT perfusion collateral index, indicating lower odds of poor cerebral collateral perfusion in males (OR 0.72, 95% CI 0.52-0.99, p=0.043). Sex was no longer significant in the sensitivity analysis (OR 0.75, 95% CI 0.55 to 1.02; p = 0.065) (Supplementary Table 4).

**Table 5.** Fractional logistic regression Odds Ratios for the CT perfusion collateral index ratio in patients for whom the volume of tissue with CBF<30% was greater than 15 mL (N=72).

|  | OR | 95%<br>confidence<br>interval | p-value |
| --- | --- | --- | --- |
| **Statin on admission** | 0.87 | 0.59 - 1.29 | 0.493 |
| **Age at onset** | 1.01 | 0.99 - 1.02 | 0.422 |
| **Sex** |  |  |  |
| Male | 0.72 | 0.52 - 0.99 | **0.043** |
| **Smoker status (reference Never smoked)** |  |  |  |
| Former smoker | 1.19 | 0.67 - 2.11 | 0.564 |
| Current smoker | 1.22 | 0.71 - 2.12 | 0.472 |
| Not answered | 1.05 | 0.74 - 1.48 | 0.798 |
| **Baseline mRS $\geq 2$** | 0.74 | 0.33 - 1.66 | 0.460 |
| **Baseline anticoagulants** | 0.92 | 0.58 - 1.45 | 0.714 |
| **Baseline antiplatelets** | 1.13 | 0.78 - 1.65 | 0.515 |
| **Diabetes** | 1.21 | 0.79 - 1.85 | 0.372 |
| **Hypercholestermia** | 1.08 | 0.73 - 1.60 | 0.709 |
| **Hypertension** | 1.02 | 0.68 - 1.53 | 0.930 |
| **Atrial Fibrillation** | 1.40 | 0.86 - 2.28 | 0.170 |

mRS: modified Rankin Score

## 7.0 Discussion

In this study we examined whether pre-stroke statin use is associated with a larger salvage volume and whether this was due to enhanced cerebral collateral perfusion. We found no relationship between statin use and salvage volume in either the total population or in the subgroup of patients that underwent EVT. Furthermore, there was no relationship between statin use and cerebral collateral perfusion. Overall, our findings indicate that the improved outcomes in stroke patients on statins prior to stroke that have been reported elsewhere are unlikely to be due to improved perfusion of the ischaemic penumbra and are likely to be primarily due to differences in initial stroke volume.

We found no significant relationship between patients being on statins prior to stroke and salvage volume. Significantly smaller infarct volume in patients on statins prior to stroke has been reported in meta-analyses and in studies with homogeneous populations [21–24]. Given the heterogeneity of our study population, the lack of significant relationship between statin use and salvage volume in our cohort is not entirely surprising. Even in our sub-analysis of a more homogenous patient cohort who underwent EVT, a procedure restricted to patients with large vessel occlusions, we found no relationship between pre-stroke statin use and salvage volume, though this was potentially influenced by the small population size (n = 86) [21,25]. There was also no relationship between TICI and salvage volume. Instead, initial infarct core volume (CBF<30%) appeared to be the main predictor of salvage volume indicating that the larger the core, the greater the potential for tissue salvage. Whilst this result may seem paradoxical, subsequent analysis indicated that a larger initial infarct core volume was strongly corelated with baseline perfusion lesion volume, which is to say that larger strokes tend to have a larger penumbra, and thus more salvageable tissue.

We also did not find a relationship between statins at admission and the CT perfusion collateral index, in line with a previous meta-analysis [23]. Several studies report no association between statin use and collateral grade [26–28], while others report that premorbid statin use is a predictor of good collaterals [16,22]. It is important to note that most of these studies measured collateral grade using semiquantitative visual scales of CTA, rather than the more quantitative CTP based perfusion collateral index, as we have done in this study.

We noted that there was no association between salvage volume and co-morbidities including diabetes, hypertension, and atrial fibrillation in our cohort. These co-morbidities are generally associated with worse stroke outcomes, larger final infarct volume, and - for diabetes and atrial fibrillation – greater infarct expansion [29–32]. Again, the studies that reported a significant relationship between outcomes and/or infarct volume examined larger cohorts or a more homogenous patient population than this study.

There are limitations to our study that should be considered when interpreting the results. First, the type of statin prescribed to patients in our cohort may have influenced the outcomes of this study. As outlined earlier, the type of statin prescribed was not listed for most patients, precluding analysis of whether some statins were associated with better outcomes than others. We also do not have data to indicate how long patients were on statins prior to stroke, how adherent they were to their dosing regimen, and what their statin dose was. Finally, our study cohort was relatively small and heterogeneous which could obfuscate effects of statins on salvage volume.

In conclusion, we aimed to determine the impact of pre-morbid statin use on salvage volume. We did not find any relationship between pre-morbid statin use and salvage volume or collateral perfusion. Instead, we found that patients with a larger stroke core volume had a greater salvage volume, indicating that the larger the core, the greater the potential for tissue salvage due to a larger initial perfusion lesion volume.

## Data Availability

The data that support the findings of this study are not openly available due to reasons of sensitivity and are available from the corresponding author upon reasonable request. Data are located in controlled access data storage through Hunter New England Health.

## 8.0 Compliance with ethical standards

### Clinical trial number

not applicable.

### Research involving human participants and/or animals

this study was conducted according to the guidelines of the Declaration of Helsinki, and approval from the Hunter New England Local Health District Human Research Ethics Committee (reference 2019/ETH03892).

### Informed consent

written informed consent was obtained from all subjects involved in the study.

## 10.0 Author’s contributions

Study conceptualisation: NJS and KGC. Data collection, curation, and compilation: KM, TL, AP. Data analysis: BT, KGC. Writing – original draft: KGC. Writing – review and editing: KGC, BT, CGE, CL, NJS. Funding acquisition: KGC, NJS.

## 11.0 Acknowledgements

We would like to acknowledge Ms. Emily Yi who assisted with compiling the data sets used in this study.

## 14.0 Glossary

Acute Ischaemic Stroke (AIS): A type of stroke caused by a sudden blockage of blood flow to a region of the brain, usually due to a clot.
Atorvastatin: A commonly prescribed statin medication used to lower cholesterol and prevent cardiovascular events.
Baseline Infarct Volume: The volume of brain tissue affected by ischaemia measured at the time of hospital admission.
Collateral Perfusion: Blood flow to ischaemic brain tissue via alternative vascular routes (collateral vessels) when primary vessels are blocked.
Computed Tomography Perfusion (CTP): An imaging technique that uses CT scanning with contrast to measure cerebral blood flow and identify ischaemic tissue.
Core (Ischaemic Core): The region of the brain that has experienced irreversible injury due to lack of blood flow.
DT6/DT2 Ratio: A quantitative index of collateral perfusion; calculated as the ratio of severely hypoperfused tissue (DT>6s) to all hypoperfused tissue (DT>2s).
Endovascular Thrombectomy (EVT): A minimally invasive procedure to remove a clot from a blocked brain artery using a catheter.
Final Infarct Volume: The volume of brain tissue that remains damaged at follow-up imaging, typically 18–36 hours post-stroke.
Hypercholesterolaemia: A condition characterised by high levels of cholesterol in the blood, increasing cardiovascular risk.
Leptomeningeal Collateral Vessels: Small vascular connections between major cerebral arteries that can provide alternative blood flow routes during arterial occlusion.
Magnetic Resonance Imaging (MRI): A non-invasive imaging technique using magnetic fields to visualise internal structures, often used to detect stroke infarct s.
Modified Rankin Scale (mRS): A functional outcome scale used to measure the degree of disability or dependence in daily activities after a stroke.
Multimodal Imaging: A combination of different imaging techniques (e.g. non-contrast CT, CTP, CTA, MRI) used to assess stroke.
National Institutes of Health Stroke Scale (NIHSS): A clinical tool used to quantify stroke severity based on neurological function.
Penumbra: The region of brain tissue surrounding the core infarct that is at risk but potentially salvageable with timely treatment.
Statins: A class of medications that reduce cholesterol production in the liver and have anti-inflammatory and vascular protective effects.
Thrombolysis in Cerebral Infarction (TICI) Score: A scale used to grade the success of blood flow restoration following endovascular therapy.

## 15.0 Supplementary data

**Supplementary Table 1.**
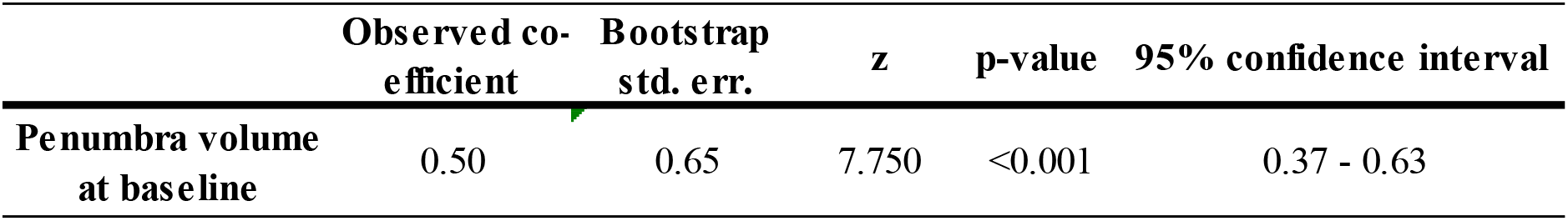
Spearman correlation of baseline penumbra volume (DT>4 minus CBF<30%) with baseline core volume (CBF<30%).

**Supplementary Table 2.** Robust regression coefficients with imputation for predictors of salvage volume (N=281).

|  | b Coefficient | 95% confidence interval | p-value |
| --- | --- | --- | --- |
| **Statin on admission** | -1.18 | -10.47 - 8.10 | 0.802 |
| **Acute core volume (mL)** | 0.93 | 0.71 - 1.15 | **<0.001** |
| **Age at onset** | -0.08 | -0.40 - 0.24 | 0.629 |
| **Sex** |  |  |  |
| Male | 3.01 | -5.04 - 11.05 | 0.462 |
| **Smoker status (reference Never smoked)** |  |  | 0.278 |
| Former smoker | -10.31 | -20.23 - -0.39 | **0.042** |
| Current smoker | -7.96 | -19.99 - 4.07 | 0.193 |
| **Baseline mRS $\geq 2$** | 10.67 | -3.13-24.46 | 0.129 |
| **Baseline anticoagulants** | 1.27 | -14.22 - 16.77 | 0.129 |
| **Baseline antiplatelets** | 1.82 | -7.25 - 10.89 | 0.693 |
| **Diabetes** | 3.00 | -6.79 - 12.80 | 0.546 |
| **Hypercholestermia** | 3.13 | -6.20 - 12.46 | 0.509 |
| **Hypertension** | 1.09 | -8.53 - 10.72 | 0.823 |
| **Atrial Fibrillation** | 3.56 | -7.25 - 14.37 | 0.517 |

mRS: modified Rankin Score; Acute core volume is measured by cerebral blood flow <30% on CTP. TICI: Thrombolysis in Cerebral Infarction. Recanalisation is defined by a Thrombolysis in Cerebral infarction (TICI) of 2c.
Imputed binary variables using logistic regression: baseline modified Rankin Score (augmented), anticoagulants use, antiplatelets use, hypercholesterinaemia, hypertension, atrial fibrillation, TICI.
Imputed categorical variables using multinomial logistic regression: Smoking status (augmented).
Other variables in the multiple imputation model conditional models: age at onset, sex, diabetes, statin on admission, acute core volume, salvage volume.

**Supplementary Table 3.** Robust regression coefficients with imputation for predictors of salvage volume in EVT subpopulation (N=95).

|  | **b Coefficient** | **95% confidence interval** | **p-value** |
| --- | --- | --- | --- |
| **Statin on admission** | -1.52 | -35.73 - 32.69 | 0.930 |
| **TICI** | 12.64 | -15.76 - 41.05 | 0.378 |
| **Acute infarct volume** | 0.48 | 0.05 - 0.92 | **0.029** |
| **Age at onset** | -0.23 | -1.28 - 0.83 | 0.668 |
| **Sex** |  |  |  |
| Male | 7.83 | -18.63 - 34.29 | 0.557 |
| **Smoker status (reference Never smoked)** |  |  |  |
| Former smoker | -25.67 | -58.56 - 7.23 | 0.124 |
| Current smoker | -4.79 | -47.69 - 38.10 | 0.823 |
| **Baseline mRS $\geq 2$** | 12.88 | -32.52 - 58.28 | 0.573 |
| **Baseline anticoagulant** | 7.32 | -39.53 - 54.18 | 0.756 |
| **Baseline antiplatelets** | 40.31 | 6.59 - 74.02 | **0.020** |
| **Diabetes** | -5.17 | -39.38 - 29.05 | 0.764 |
| **Hypercholestermia** | -12.50 | -46.28 - 21.27 | 0.462 |
| **Hypertension** | 15.78 | -17.06 - 48.62 | 0.342 |
| **Atrial Fibrillation** | 7.17 | -34.31 - 48.65 | 0.732 |

Acute core volume is measured by cerebral blood flow <30% on CTP; mRS: modified Rankin Score; TICI: Thrombolysis in Cerebral Infarction. Recanalisation is defined by a Thrombolysis in Cerebral infarction (TICI) of 2c. Imputed binary variables using logistic regression: Baseline modified Rankin Score (augmented), presence of atrial fibrillation, TICI binary. Imputed categorical variables using augmented multinomial logistic regression: Smoking status (augmented). Other variables in the multiple imputation model conditional models: age at onset, sex, anticoagulants use, antiplatelets use, hypercholesterinaemia, hypertension, diabetes, statin on admission, acute core volume, salvage volume.

**Supplementary Table 4.** Fractional logistic regression Odds Ratios with imputation for the CT perfusion collateral index in patients for whom the volume of tissue with CBF<30% was greater than 15 mL (N=72).

|  | **b Coefficient** | **95%**<br>**confidence**<br>**interval** | **p-value** |
| --- | --- | --- | --- |
| **Statin on admission** | 0.88 | 0.59 - 1.32 | 0.536 |
| **Age at onset** | 1.01 | 0.99 - 1.02 | 0.358 |
| **Sex** |  |  |  |
| Male | 0.75 | 0.55 - 1.02 | 0.065 |
| **Smoker status (reference Never smoked)** |  |  |  |
| Former smoker | 1.15 | 0.69 - 1.92 | 0.598 |
| Current smoker | 1.30 | 0.78 - 2.17 | 0.320 |
| **Baseline mRS $\leq 2$** | 0.68 | 0.32 - 1.46 | 0.328 |
| **Baseline anticoagulants** | 0.92 | 0.60 - 1.39 | 0.688 |
| **Baseline antiplatelets** | 1.12 | 0.79 - 1.60 | 0.516 |
| **Diabetes** | 1.20 | 0.79 - 1.81 | 0.388 |
| **Hypercholestermia** | 1.06 | 0.72 - 1.58 | 0.759 |
| **Hypertension** | 1.06 | 0.70 - 1.62 | 0.772 |
| **Atrial Fibrillation** | 1.42 | 0.90 - 2.24 | 0.129 |

mRS: modified Rankin Score

